# Large language model-based evaluation of the impact of gender in medical research

**DOI:** 10.64898/2026.01.06.26343564

**Authors:** Michael S. Yao

**Affiliations:** University of Pennsylvania, Philadelphia, PA 19104

## Abstract

**Background:** Gender disparities in academic medicine have been previously reported, but prior bibliometric studies have been limited by small sample sizes and reliance on manual gender annotation methods. These bottlenecks constrain previous analyses to only a small subset of clinical literature. To assess gender-based differences in authorship trends, research impact, and scholarly output over time in clinical research at scale, we hypothesized that large language models (LLMs) can be an effective tool to facilitate systematic bibliometric analysis of academic research trends.

**Methods:** We conducted a retrospective, cross-sectional bibliometric study evaluating manuscripts published between January 2015 and September 2025 across over 1,000 PubMed-indexed academic medical journals. Over 1 million manuscripts, written by more than 10 million authors across 13 medical specialties, were analyzed. To enable this large-scale study, the genders of manuscript authors were annotated using a scalable LLM-based pipeline compatible with consumer-grade hardware.

**Results:** We found that the proportion of female principal investigators has increased over time across different medical subspecialties. However, studies led by male authors tended to be published in higher-impact journals and cited more frequently than those led by female authors. We also observed that researchers of the same gender tended to work together when compared to colleagues of the opposite gender.

**Conclusions:** While our findings revealed persistent gender-based differences in authorship trends, citation practices, and journal placement, we also observed ongoing, meaningful progress in female representation within academic medical research over time. Our results suggest that LLMs can be a powerful tool to scalably and periodically track this continued progress in future academic medical research.

**Plain Language Summary:** Academic research is important to advance the field and practice of medicine. To obtain an accurate picture of the differences in medical research and impact between male and female researchers, we leveraged large language models (LLMs) to identify author genders for over one million medical research papers published between 2015 and 2025. We found that the number of women serving as lead researchers has increased over time across many medical specialties. However, important gaps in achieving gender equality in medical research remain. Our study ultimately helps demonstrate that LLMs can help us monitor gender-based trends in academic research in the future.

## Introduction

Gender disparities in academic medicine have been extensively documented, with evidence of persistent inequities in authorship, leadership, and recognition across multiple clinical specialties.^1–4^ Prior studies have reported that women remain underrepresented in both first and senior authorship roles across disciplines, with implications for academic advancement and visibility, citation patterns, and grant funding.^5–9^ However, these quantitative investigations in medicine have often relied on manual annotation using data from a small set of journals that do not fully capture the breadth of medical literature (**Supplementary Table 1**). This limits our ability to detect large-scale and nuanced trends across academic medicine and its subspecialties.

Understanding the scope and evolution of gender disparities in academic research is crucial to promote equity and guide institutional and policy interventions. Prior work has demonstrated that inequities in authorship are associated with differences in citation counts, journal placement, and overall scholarly impact.^1,10–12^ Furthermore, the dynamics of authorship composition—such as homogeneity of gender within research teams and how gender may correlate with the downstream impact of research—have not been systematically studied.

Recent work on large language models (LLMs) offers a new opportunity to address these limitations.^13,14^ By learning from large corpora of natural language, LLMs have demonstrated strong performance in data extraction and classification tasks, including annotation of structured information from unstructured text.^15–17^ Based on these findings, we hypothesize that LLMs may be able to help automate the labor-intensive task of author gender classification in bibliometric analyses, enabling scalable evaluation across large corpora of manuscripts. Such methods can reduce reliance on manual curation, mitigate individual annotator bias, and facilitate frequent monitoring of trends over time. However, the potential of LLMs to systematically examine gender disparities in medical research has not yet been rigorously explored.

The purpose of this study is to investigate how LLMs can be used to conduct a large-scale, retrospective bibliometric analysis of academic medical literature. Specifically, we report on quantitative metrics of authorship trends and research impact stratified by author gender across a decade of publications from 2015 to 2025. We then compare our findings with those of prior studies to better characterize the validity of our LLM-based approach. By leveraging our proposed gender annotation method using LLMs, we provide new insights into both persistent inequities and emerging shifts in medical scholarship.

## Materials and Methods

This retrospective study was determined to be not human subjects research because it exclusively used publicly available bibliometric data from PubMed, and did not involve human subjects, patient data, identifiable private information. Therefore, Institutional Review Board approval of this research was not required per the Common Rule (U.S. Department of Health and Human Services, 45 CFR 46).

### Journal and Manuscript Data Curation

We first sought to determine the set of academic journals whose manuscripts would be included in our study. To this end, we used the Entrez programming utilities application programming interface (API) to query the National Library of Medicine (NLM) Catalog database for journals that are both (1) published in English; and (2) labeled by an NLM-designated Broad Subject Term related to a “medical specialty,” which refers to either general medicine or a medical subspecialty.^18^ See **Supplementary Table 2** for additional details. Journals that stopped publishing manuscripts prior to 2015 were excluded.

Using the same Entrez API and a custom Python script, we then queried the PubMed Central (PMC) database for manuscripts published in an NLM-catalogued journal identified above. For each manuscript, we used the API to fetch its author list, year of publication, and digital object identifier (DOI). We restricted our analysis to manuscripts published between January 2015 and September 2025 (**Fig. 1a**).

**Figure 1:**
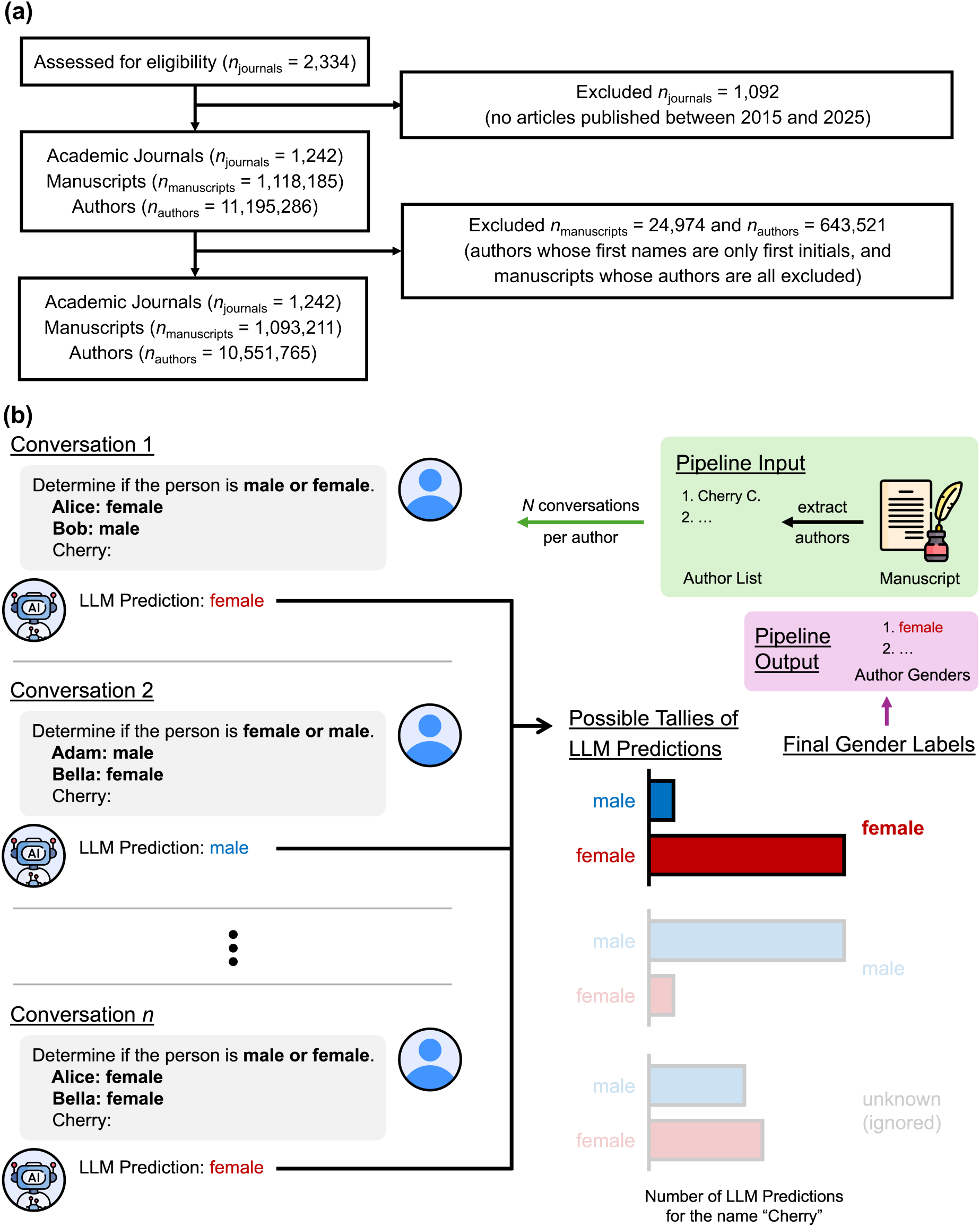
Study flowchart and method overview. **(A)** Journal, manuscript, and author data were sourced from the National Library of Medicine (NLM) Catalog and PubMed Central (PMC); all manuscript data published between January 2015 and September 2025 were included in our analysis. We excluded authors whose first names were only limited to their initials, narrowing the scope of our bibliometric analysis to 1,242 unique academic medical journals, 1,093,211 unique manuscripts, and 10,551,765 authors. **(B)** For each author of a research manuscript, we query a large language model (LLM) to predict the author’s gender from their first name given different semantically equivalent input prompts. Names that were consistently labelled “female” or “male” under different prompt variations were assigned the corresponding gender label.

To quantitatively measure a journal’s academic impact, we used the SCImago Journal Rank (SJR) published by the SCImago Research Group, which measures a journal’s citation impact over a 3-year period, where higher is better.^19^ For each manuscript published in a journal, we labelled a manuscript entry with the journal’s SJR from the year immediately prior to the publication year of the manuscript. The SJR metric can therefore be thought of as a proxy for the impact of an academic journal. We also recorded the Open Access (OA) status of each journal, which was also publicly provided by the SCImago Research Group; briefly, an OA journal generally publishes scholarly articles online for free, immediate access.

Finally, we wrote a separate custom Python script to query the OpenCitations API (v1.2.0) to obtain a manuscript’s total number of citations; number of “self-citations” (defined as a citation from a separate manuscript that shares at least one author, which is tracked in the OpenCitations database); and total number of references.^20^

### Predicting the Gender of Manuscript Authors Using a Large Language Model

Prior works analyzing smaller sets of academic manuscripts have almost exclusively used human annotators or fixed-size databases to manually label the gender of manuscript authors.^2,3,5–7,21^ However, such efforts scale poorly and are sensitive to potential internal biases of the annotators.^22^ Recent work has looked to that large language models (LLMs)—which are generative autoregressive models trained on large corpora of text—to effectively automate data annotation pipelines.^23^ Here, we leverage an LLM-based gender annotation approach inspired by contemporary research on leveraging probabilistic sampling and prompt invariance as proxies for the trustworthiness of an LLM’s output.^24–29^ We hypothesize that an accurate LLM gender prediction should be robust to semantically insignificant changes to the input prompt (**Fig. 1b**). For a given author’s first name 𝑥*_s_*, we use 𝑛 = 5 different input prompts that phrase the same request using different words to query the Llama-3.1 8B Instruct model from Meta AI (meta-llama/Llama-3.1-8B-Instruct) to predict the gender of the author. This process gives us a set of 𝑛 independently sampled predictions 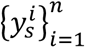, where each 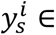 {male, female}. Using a threshold value of 𝜏 = 4, we then compute the final label 𝑦_s_ according to

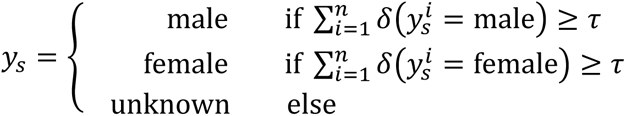

where 𝛿 is the delta function. Put simply, the final prediction was “male” or “female” if at least 4 out of the 5 LLM predictions 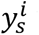 were consistent with one another; otherwise, the final prediction was labeled as “unknown.” Our chosen threshold is consistent with prior work.^30^ We use an LLM temperature parameter of 0.01 for each prediction step; see the **Supplementary Methods** and **Supplementary Results** for additional details.

To validate this LLM-based approach, 2 human annotators individually completed 2 hours of implicit bias training and were then tasked with manually annotating the gender of a random subset of 1000 authors sampled across all 13 medical specialties. Each first name was annotated with exactly one of the following labels: “male,” “female,” or “unknown,” where “unknown” corresponds to cases of either unisex first names or when the annotator could not determine the gender. The annotators were only given access to author first names and could use any online tools to complete the task. We then compared the predicted labels 𝑦_s_ from the described LLM-based pipeline with the corresponding reference human annotations.

### Statistical Analysis

To account for multiple comparisons, we used a Bonferroni-corrected significance threshold of *P <* 0.001 to indicate a statistically significant difference. *P* values less than 2.225 x 10^-308^ (i.e., the smallest 64-bit normal float that can be represented in IEEE-754 format) are reported as *P* = 0.0. All statistical analyses were performed using Python software (version 3.13.7; Python Software Foundation) and the SciPy scientific computing package (version 1.16.1; NumFOCUS). Data deduplication was performed based on unique identifiers (NLM IDs for journals, PMC IDs for articles, and DOIs for citation analysis) to prevent artificial inflation of sample sizes. All experiments were run on one 96-core Intel Xeon CPU and one NVIDIA RTX A6000 GPU (CUDA version 12.6).

## Results

### Data Description

We identified a total of 2,334 unique journals in the NLM Catalog and filtered them to the subset of 1,242 journals that published at least one manuscript between January 2015 and September 2025 (**Fig. 1a**). From these journals, approximately 11 million manuscript authors were identified from PubMed Central. We then excluded approximately 650 thousand (5.75%) authors because their first names were only initials (making it impossible to reliably determine their gender). Any manuscripts whose authors were all excluded due to the above criteria were also excluded. Our final dataset included 1,093,211 manuscripts and 10,551,765 authors publishing in 1,242 unique academic journals. Additional analysis of the distribution of research articles published within each journal are included in **Supplementary Fig. 1**, and the full dataset of medical research journals included in our study is detailed in **Supplementary Data 2**.

### Validation of Our LLM Gender Annotation Pipeline

The two human annotators achieved a Cohen’s kappa coefficient of κ = 0.945, indicating strong inter-rater reliability in manually labelling the gender of authors by first name. Restricting our analysis to the subset of first names where the two human annotators agreed on the final gender label, we found that our proposed LLM-based method achieved a Cohen’s kappa coefficient of κ = 0.967 with respect to the consensus labels between the human annotators. Furthermore, our approach also achieved an overall accuracy of 98.6%, and an accuracy of 98.5% (resp., 98.6%) on predicting the gender of female (resp., male) authors when compared to the reference human consensus labels. We compare our proposed LLM-based gender annotation strategy against alternative approaches in **Supplementary Tables 1 and 3**.

We also compared the gender annotation rate of our LLM-based approach to that of other gender annotation methods used in prior works. We define the gender annotation rate as the proportion of manuscript authors that were able to be assigned a gender of either “male” or “female.” Our method achieved an annotation rate of 85.2% across all specialties, which is comparable to prior work using database-based approaches for gender annotation.^4,30,31^ Other works that use manual, human-based annotation methods have been able to achieve higher annotation rates as high as 98.5%;^2,32,33^ however, these prior studies also acknowledge that the success of their methods relied on the ability of researchers to search for the gender of the author online based on their public Internet profile. In contrast, web search capabilities were not made available to our LLM-based method.

### Manuscript Authorship Over Time

To better characterize the genders of manuscript authors using the consensus gender labels predicted by our proposed method, we first computed the fractions of male and female manuscript authors. We found that the proportion of male authors was greater than that of female authors for all evaluated medical specialties except for Primary Care and Geriatrics (**Fig. 2**). However, when focusing on only the first authors of each manuscript, female gender representation was greater in 8 out of the 13 specialties (General Medicine, Allergy, Endocrinology, Primary Care, Geriatrics, Oncology, and Rheumatology). The last authors of manuscripts were more frequently male than female across all medical subspecialties. Traditionally, the first author is typically the researcher who leads the research work, while the last author is the Principal Investigator (PI) responsible for the overall integrity of the work.

**Figure 2:**
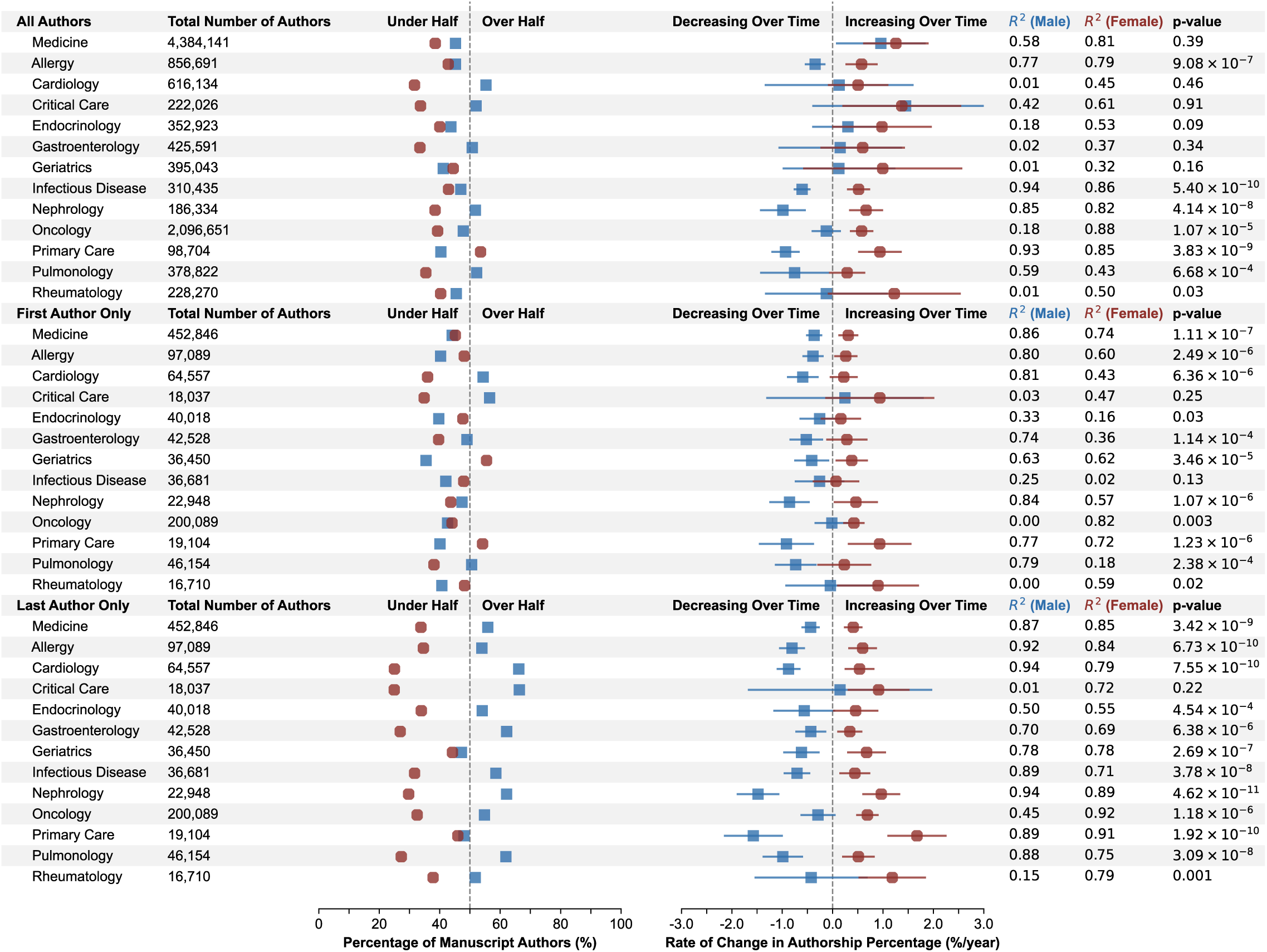
Gender analysis of manuscript authors and authorship trends. Within each of the 13 medical subjects defined by the National Library of Medicine (NLM), we show a scatter plot of the proportion of (**top**) all manuscript authors; (**middle**) first authors; and (**bottom**) last authors that are predicted to be male (**blue squares**) or female (**red circles**). We also fit separate linear regression models to describe how these gender proportions change as a function of publication year. We then plot the estimated rate of change in authorship percentage and report the coefficient of determination (*R^2^*) of each regression model; error bars represent 99.9% confidence intervals of the true parameter values. Finally, we conducted two-sided, unpaired Wald tests with the Satterthwaite approximation to determine if the rates of change are different between male and female authors within each row; the corresponding *p*-values are listed in the final column.

We then sought to determine if these gender proportions have changed over time. To this end, we estimated separate ordinary least squares (OLS) regression models to predict the observed proportion of each gender each year within each subspecialty (**Fig. 2**). Broadly, we found that female gender representation was increasing over time with statistical significance in the population of all manuscript authors for 8 out of 13 medical subspecialties; first authors for 6 subspecialties; and last authors for 12 subspecialties. Notably, these results are consistent with prior work describing increasing female representation as PIs of academic research labs.^34,35^ Additional quantitative results are shown in **Supplementary Fig. 2**.

### Within-Manuscript Gender Distribution of Author Lists

We next examined the distribution of proportions of female and male authors within author lists of a given manuscript (**Figure 3**). Understanding patterns of gender clustering within individual manuscript author lists may help elucidate whether researchers tend to publish and work with other researchers of the same (or different) genders. Restricting our attention to manuscripts with at least two authors, we found that research teams tended to fall into one of three configurations: primarily male, primarily female, or mixed gender with roughly equal representation of male and female researchers. Qualitatively, we observed that manuscripts were more likely to be primarily male than primarily female in Medicine, Cardiology, Critical Care, Gastroenterology, Infectious Disease, Nephrology, Oncology, and Pulmonology. In contrast, we observed the opposite trend in the fields of Geriatrics and Primary Care.

**Figure 3:**
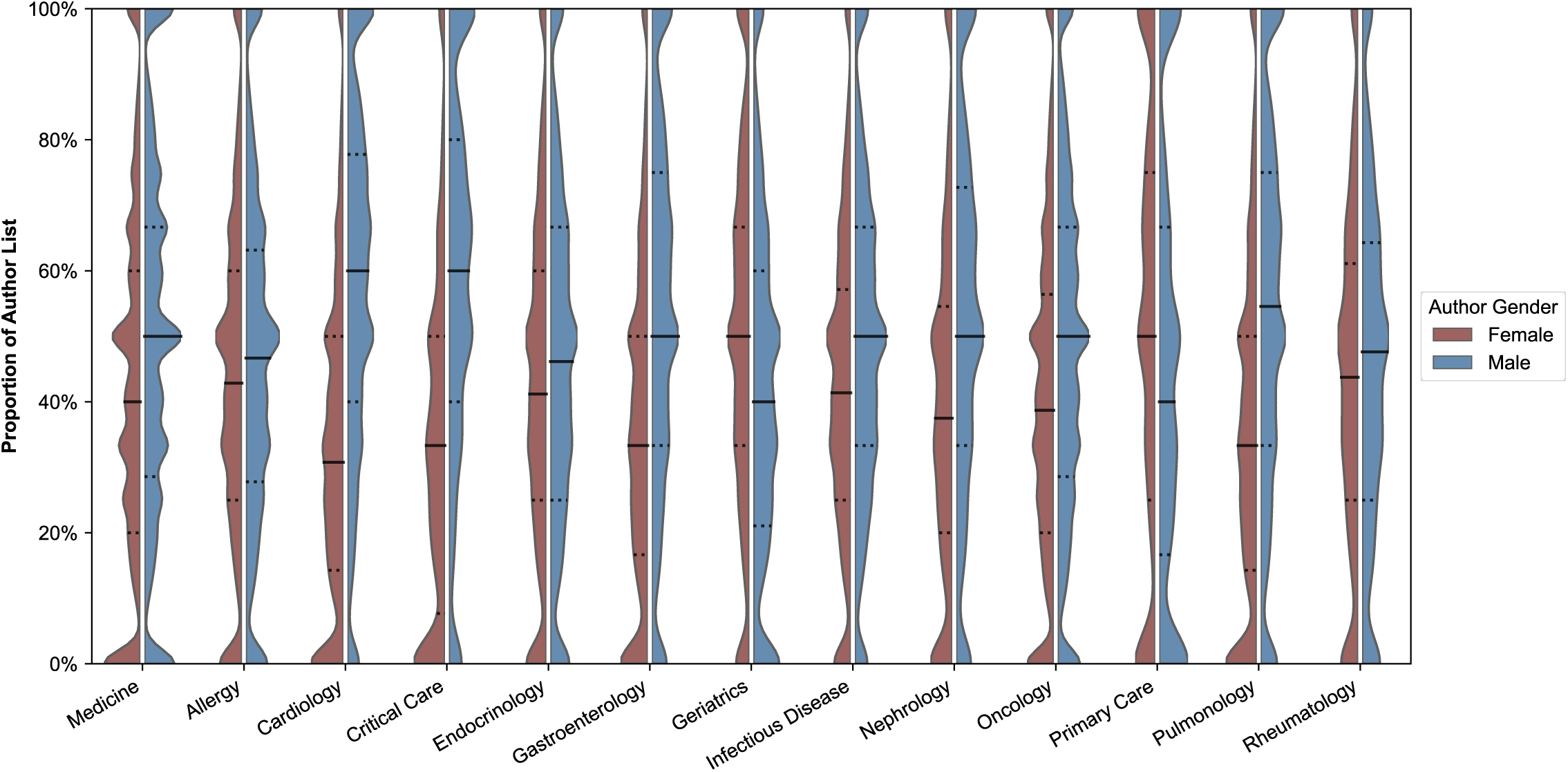
Analysis of gender distributions within manuscript author lists. For each medical subjects defined by the National Library of Medicine (NLM), we plot the distribution over manuscripts of the percentage of author lists that are predicted to be female (red left side of each violin plot) and male (blue right side of each violin plot). The median of each distribution is indicated by solid black lines, and the first and third quartiles are indicated by dotted black lines.

Within each medical specialty, we then ran a *z*-test for intra-group homophily under a random assignment model to quantitatively assess whether authors of the same gender tended to work together more frequently than what would be expected due to chance. The *p*-values across all specialties were each individually less than 10^-112^, suggesting a statistically significant trend consistent across all medical specialties that researchers of the same gender tend to work together as opposed to with researchers of the opposite gender. We found that the odds ratio (OR) of last authors working with a first author of the same gender as opposed to the opposite gender was 2.117 ± 0.031 (mean ± 99.9% CI) averaged across all specialties. Additional analysis included in **Supplementary Table 5** and our **Supplemental Results**. Finally, we also analyze the gender-based distribution of single-author manuscripts in each medical discipline in **Supplementary Table 8**.

### Manuscript Citation and Reference Counts

Citations and references constitute key indicators of scholarly impact.^5–9^ A citation refers to an instance where the manuscript is cited by a subsequent publication, whereas a reference is a previously published external work cited within the manuscript. We examined whether these metrics varied according to the gender of the first or last author. In **Figure 4**, we found that manuscripts with male first or last authors generally demonstrated higher citation counts than those authored by females in corresponding positions, with statistically significant gender differences observed in 8 of the 13 evaluated medical subspecialties for first authors and 10 subspecialties for last authors. Additionally, male last authors were associated with a greater number of “self-citations” per manuscript, which were defined as citations originating from publications sharing at least one author with the cited work. Although differences in reference counts between genders were occasionally statistically significant, we found that the absolute magnitude of these differences was relatively small: the estimated gender-based difference in the number of references averaged across all subspecialties was less than 1 reference for both the first and last author groups.

**Figure 4:**
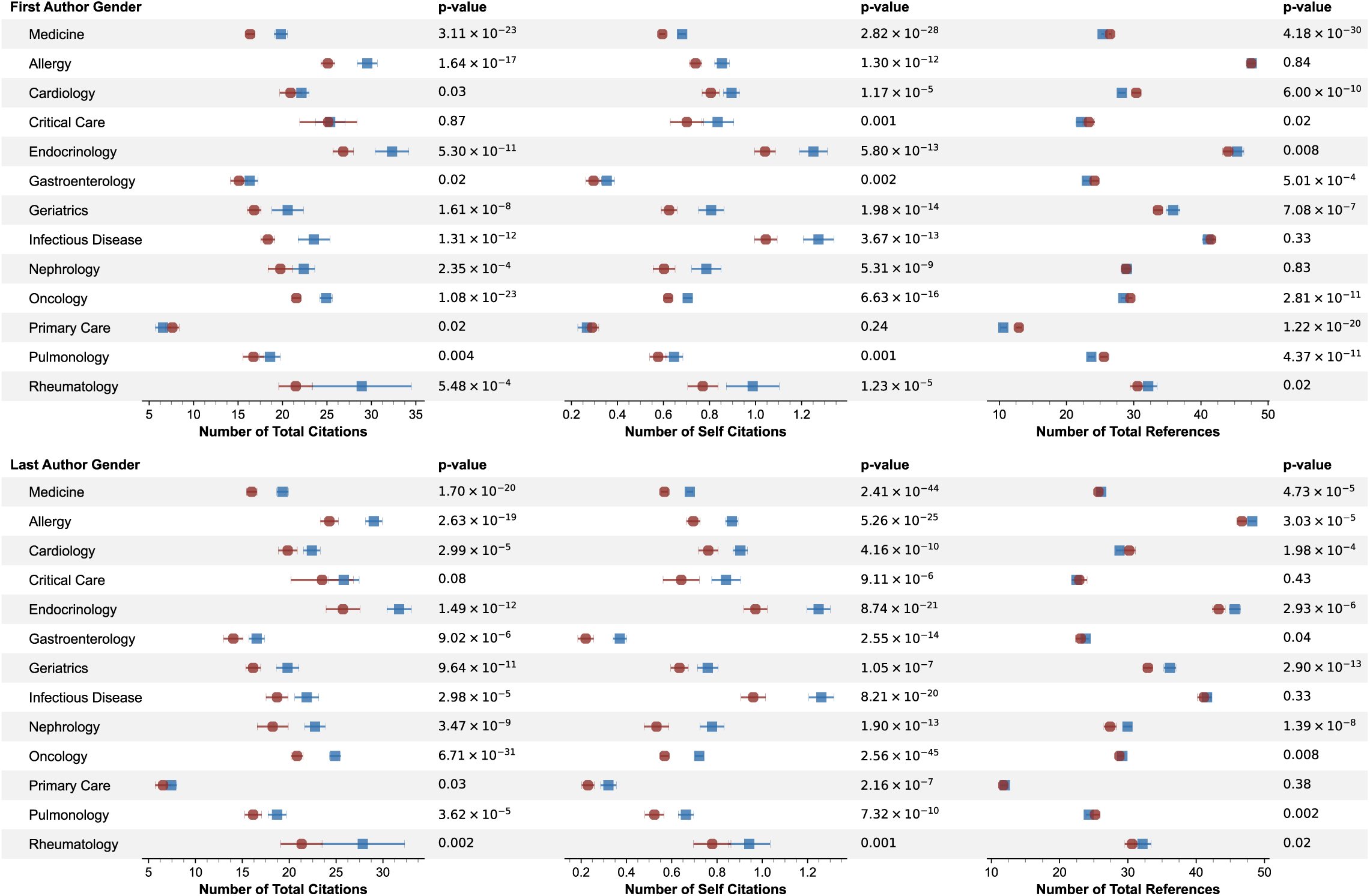
Manuscript citation and reference trends as a function of author gender. We plot the mean and 99.9% confidence intervals of the number of (**left**) total citations; (**middle**) self-citations; and (**right**) total references as a function of the gender of the first and last author. For each medical subspecialty and metric, we also ran a two-sided unpaired *t*-test to determine if the values of each metric are significantly different between first and last authors of different genders within each row; the corresponding *p*-values are listed in the final column.

### Publication Journal Impact and Open Access Status

Finally, we examined the SCImago Journal Rank (SJR) of the journals that published articles written by first and last authors of different genders across different medical subspecialties. We observed a statistically significant difference between the mean SJR of journals publishing papers written by male and female first (resp., last) authors in 11 (resp., 10) out of 13 subspecialties (**Fig. 5**). However, the absolute difference in mean SJR was relatively small in most cases; the average SJR associated with manuscripts published by male last authors was greater than that of female last authors by an absolute [relative] difference of 0.31 points [9.46%], 0.27 points [12.8%], 0.20 points [7.59%], and 0.20 points [10.6%] in the fields of Critical Care, Endocrinology, Allergy, and Nephrology, respectively. The observed mean gender-based difference in SJR was also greater as a function of the last author’s gender than as a function of the first author’s gender in all evaluated medical specialties except for Pulmonology. This is expected, as the last author (i.e., the Principal Investigator) is typically responsible for evaluating the suitability of a manuscript for a particular journal.

**Figure 5:**
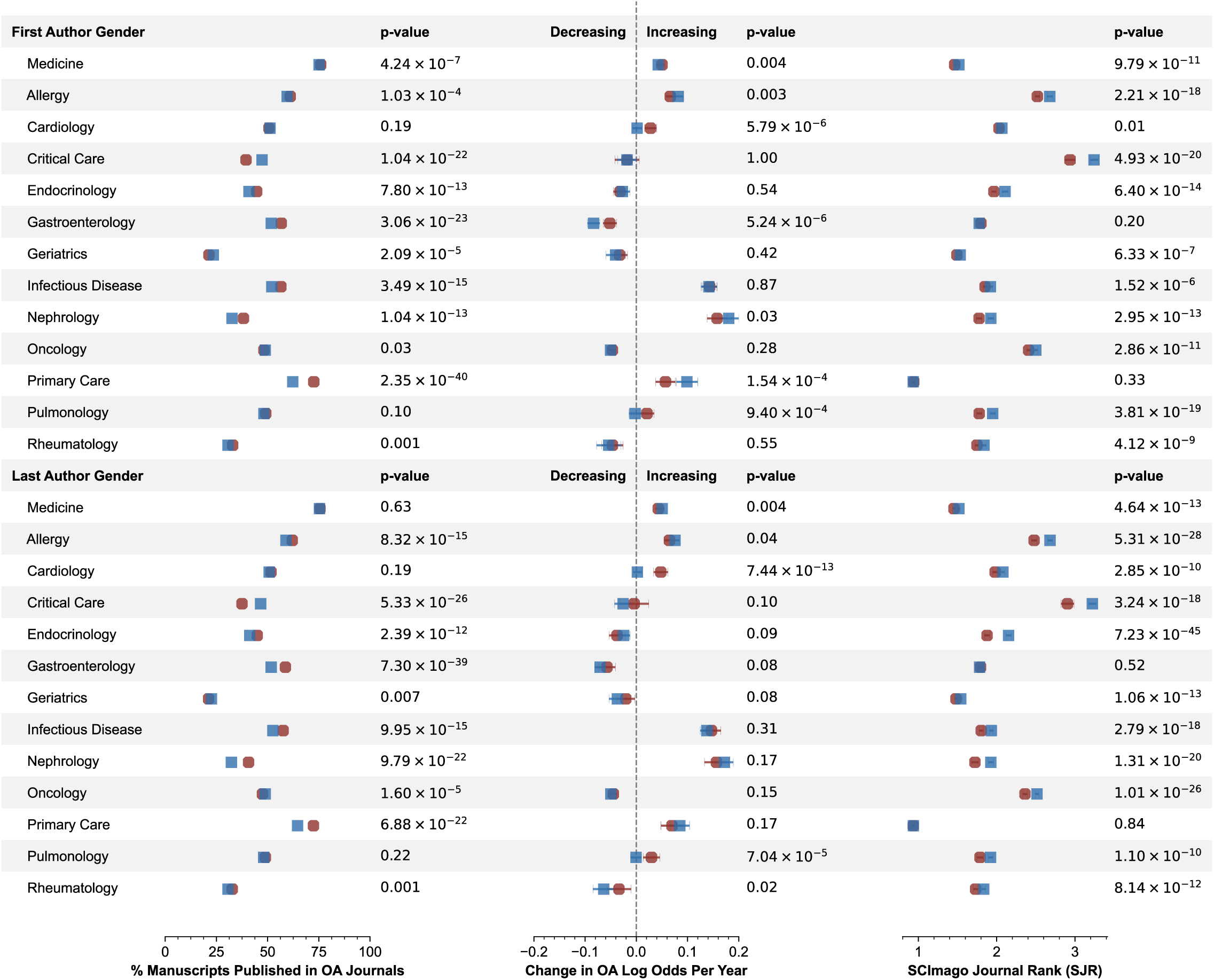
Gender-based differences in SCImago Journal Rank (SJR) and journal Open Access (OA) status. (**Left**) For each medical specialty, we first plot the percentage of manuscripts published in OA journals as a function of the gender of the first and last authors. The *p*-values are computed from a two-sample *z*-test to compare the time-averaged marginal effect of gender on the year-over-year proportion of OA manuscripts. (**Middle**) We then fit separate logistic regression models for each specialty and author position to estimate the probability of a manuscript being published in an OA journal as a function of author gender and year, and plot the mean and 99.9% confidence interval of the change in OA log odds per year by gender according to the fitted model. The reported *p*-values are computed from a two-sample, unpaired *t*-test to compare the difference in mean change in OA log odds per year by gender. (**Right**) Finally, we plot the mean and 99.9% confidence interval of the journal SJR as a function of author gender. The reported *p*-values are computed from a two-sample, unpaired *t*-test to compare the difference in mean SJR by gender.

We also sought to evaluate the proportion of manuscripts published in Open Access (OA) journals as a function of publication year and gender. To this end, we estimated a generalized linear model with a binomial outcome using a logit link, with the proportion of OA manuscripts within each discipline as the dependent variable and gender, year, and their interaction as predictors. Our results suggest that manuscripts authored by female first or last authors were more likely to be published in OA journals when compared to their male counterparts in the fields of Gastroenterology, Infectious Disease, and Primary Care. In contrast, male first and last authors in Critical Care published more frequently in OA journals than female first and last authors, respectively. While there were some instances of subtle gender-based differences in the change in the proportion of OA manuscripts year over year, the differences by medical specialty were much more pronounced: the fields of Infectious Disease, Nephrology, and Primary Care were associated with the greatest increases in the proportion of OA manuscripts per year regardless of the genders of the first and last authors.

## Discussion

Our study contributes to the growing body of literature examining gender disparities within medical research. Previous investigations have largely relied on manual annotation and were constrained by relatively small sample sizes, limiting their ability to capture large-scale trends (**Supplementary Table 1**).^21,7,6,5,8^ To this end, our findings demonstrate that LLMs can help scale the systematic evaluation of authorship and citation patterns in academic medicine, enabling broader and more nuanced insights into issues of gender representation in the field.

First, consistent with prior work, clinical and medical research led by male senior PIs continue to receive more citations than those led by female senior PIs.^2,3,9^ Furthermore, the overall percentage of female first authors and senior authors has been only slowly increasing by an average of approximately 1% per year over the past decade (**Fig. 2**). Our analysis also reveals structural imbalances in team composition, with many authorship groups exhibiting strong gender homogeneity rather than gender-diverse collaborations (**Figure 3**). Collectively, these findings underscore ongoing disparities in authorship recognition and research practices in academic medicine.

Taken together, our results highlight both persistent inequities and promising shifts in medical research by gender. Continued structural and cultural interventions are needed to promote equity in authorship, collaboration, and scientific recognition. Importantly, our study also reproduces key findings from prior works analyzing smaller, domain-specific subsets of manuscripts, strengthening the validity of our LLM-based approach for gender-based bibliometric studies.^2,4,30,34,36^ Our results ultimately demonstrate the utility of LLM-driven analyses for ongoing monitoring of research trends. Future work might examine how to extend our method to evaluate the longitudinal effects of policy initiatives and identify mechanisms that promote equitable participation in medical research.

There are important limitations associated with our work that warrant discussion. A core step of our experimental pipeline was using language models to predict the genders of authors based on their first name. However, individual gender identities may not align with LLM-generated predictions. Furthermore, recall that we used the Llama-3.1 8B model from Meta AI (a US-based company) to automate the annotation of gender labels by author first name. We chose to use this particular language model because it is easily accessible on consumer-grade hardware and performed well on the task at hand. However, it may also underperform on authors with first names that are not common in the United States. For example, Chinese first names written in Mandarin often encode gender in the specific Chinese characters used, which is important information that is lost when the name is translated to English. Separately, different subpopulations of authors may have different incidences of unisex names or choose to publish using only first initials. Further discussion of the observed failure modes associated with our LLM-based gender annotation pipeline is included in **Supplementary Table 4**. As a result, certain subpopulations of authors may have been systematically excluded from our analysis. Future work might explore how to use modern multilingual and more performant language models to better close this potential gap.^37,38^

We also note that our current method is not designed to recognize non-binary and gender-diverse individuals who may not identify with the gender most commonly associated with a first name at a population level.^39^ Without additional author metadata catalogued at a systemic level, it is challenging to factor in these nuances into our proposed method.

Furthermore, other variables of interest, such as author nation of origin and institutional affiliation, might be interesting to explore using a similar LLM-based method in follow-up bibliometric analyses. However, a key challenge with this proposed study is in scalably and accurately determining these variables for large groups of manuscript authors, as prior work has relied on human efforts to laboriously query the Internet for this information on an author-by-author basis. However, recent advancements in the web-search and multi-step reasoning capabilities of language models may help address this limitation.^40–42^ We leave a rigorous exploration of how to best integrate these technologies into bibliometric analyses as an opportunity for future work.

We also limited our analysis to research available on PubMed, which only contains a subset of the total body of medical research. For example, non-archival materials including conference abstracts, oral presentations, and research posters are typically not deposited in PubMed. Some research teams may also choose not to openly publish their research to protect intellectual property, preserve competitive industry know-how, or a variety of other reasons. Our method is also inherently tied to the accuracy and maintenance of external, third-party resources such as the OpenCitations and PMC databases. The manuscript coverage of these repositories may be incomplete and vary across both academic fields and time, which may impact the validity of our results. Future work may be warranted to better characterize the real-world significance of this potential limitation.

## Supporting information

Supplementary Data Descriptions

## Abbreviations

API: application programming interface
DOI: digital object identifier
LLM: large language model
NLM: National Library of Medicine
OA: open access
OLS: ordinary least squares
PMC: PubMed Central
SJR: SCImago Journal Rank

## Data Availability

All data generated or analyzed during the study are included in the published paper, and also at the following URL: https://huggingface.co/datasets/michaelsyao/MedicineAuthorship.

## Code Availability

We make our custom code implementation publicly available at https://github.com/michael-s-yao/medicine-authorship.

## Acknowledgements

The authors thank Allison Chae at Main Line Health for their help with the empirical evaluation of our LLM-based gender annotation pipeline. We also appreciate the computational resources made available by Osbert Bastani at the University of Pennsylvania to support this work. M.S.Y. was supported by the NIH (F30 MD020264) and research grants from OpenAI and GenderAPI (Ozan Soft). The content is solely the responsibility of the authors and does not necessarily represent the official views of the NIH.

## Notes

### Competing Interest Statement

The authors have declared no competing interest.

### Funding Statement

This study was supported by the NIH (F30 MD020264) and research grants from OpenAI and GenderAPI (Ozan Soft).

