## Supplementary Information for "Large language model-based evaluation of the impact of gender in medical research"

Michael S. Yao[1]

**Affiliations**

[1]University of Pennsylvania, Philadelphia, PA 19104

**Corresponding Author:**

Dr. Michael S. Yao
University of Pennsylvania
3330 Walnut St
Philadelphia, PA 19104


**Table of Contents**

Supplementary Figures 1 – 2 …… S1 – S2
Supplementary Tables 1 – 8 …… S3 – S10
Supplementary Methods …… S11
Supplementary Results …… S12 – S14
Supplementary References …… S15

#### Supplementary Figures

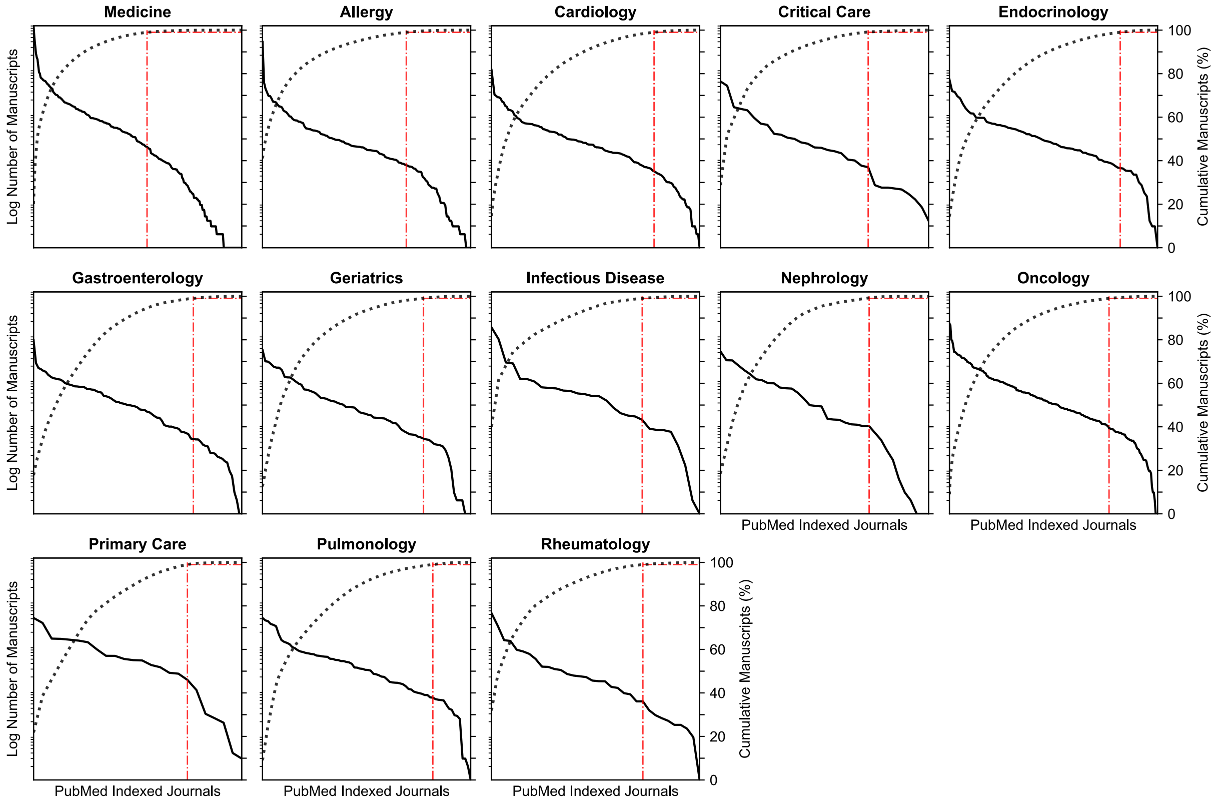


**Supplementary Figure 1: Distribution of research articles by journal.** For each medical specialty, we plot a Pareto chart of the PubMed Indexed Journals on the *x*-axis sorted by the number of manuscripts published in each journal between January 2015 and September 2025, plotted on the left *y*-axis in solid black. We also plot the cumulative proportion of published manuscripts in each specialty in dotted dark grey on the right *y*-axis. The fraction of journals that contain at least 99% of published manuscripts in the specialty is shown by the dotted-dashed red line. Note that all journals within each specialty have been compressed down to the same *x*-axis "length" – the total absolute count of journals shown varies by specialty.

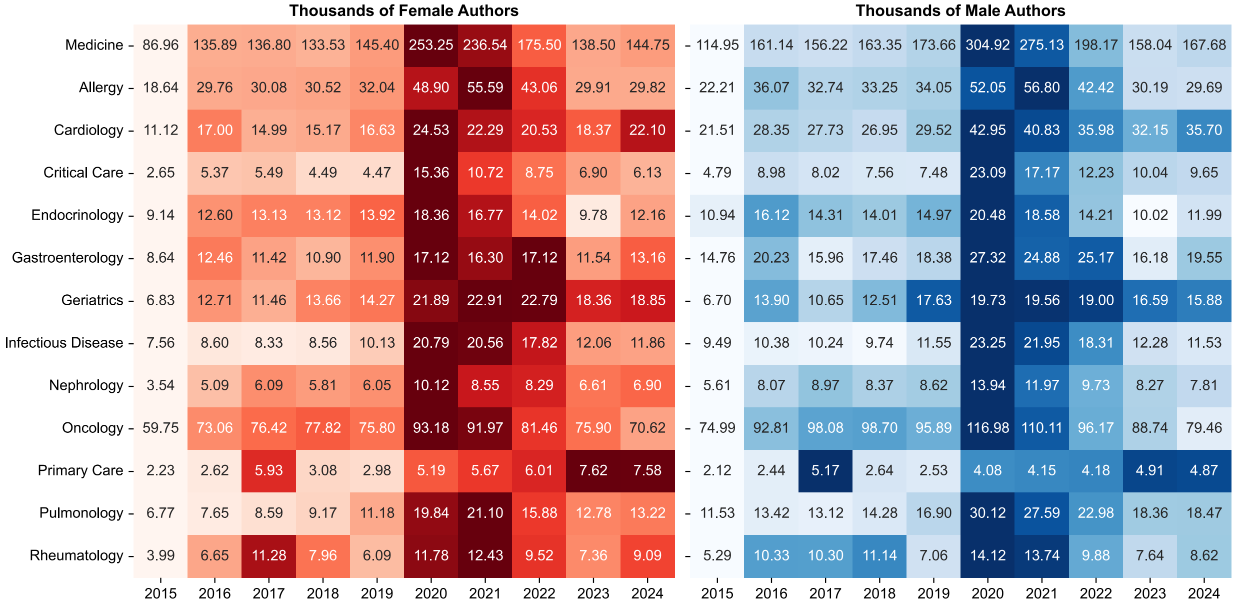


**Supplementary Figure 2: Number of authors of medical research papers over time.** The left (resp., right) heatmap shows the number of female (resp., male) authors of research papers in each medical specialty (rows) by year (columns). Note that while our main study considers manuscripts published between January 2015 and September 2025, we do not show the number of authors from the 2025 calendar year, which was incomplete at the time that our study was conducted.

#### Supplementary Tables

| Publication | Number of Authors | Number of Manuscripts | Number of Journals | Gender Annotation Strategy | Time Period | Annotation Success Rate |
|---|---|---|---|---|---|---|
| Bruns N et al. Crit Care (2025).[1] | 77,713 | 42,970 | 5 | Proprietary (NamSor) | 2005-2024 | 87.3% |
| Brück O. Nat Commun Med (2023).[2] | 4,217 | 10,058 | 5 | Database (GenderizeR) | 2010-2019 | 94.1% |
| Böhme K et al. Ped Res (2022).[3] | 363,518 | 156,642 | 400 | Database (Gendermetrics.Net) | 2008-2018 | 76.0% |
| Krstacic JE, et al. PLoS One (2022).[4] | 2,900 | 1,080 | 3 | Manual | 2012-2019 | 98.4% |
| Yao J, et al. J Glob Health Rep (2022).[5] | 190,809 | 33,854 | 46 | Database (Gender-API) | 1945-2020 | 74.8% |
| Chatterjee P, et al. JAMA Netw Open (2021).[6] | 10,494 | 5,554 | 5 | Database (GenderizeR) and Manual | 2015-2018 | 98.1% |
| Campbell JC, et al. J Am Coll Radiol (2019).[7] | 11,657 | 1,934 | 3 | Manual | 2011-2015 | N/A |
| Jagsi R et al. N Engl J Med (2006).[8] | 7,249 | 3,872 | 6 | Manual | 1970, 1980, 1990, 2000, and 2004 | 98.5% |
| **Ours** | 10,551,765 | 1,093,211 | 1,242 | LLM | 2015-2025 | 85.2% |

**Supplementary Table 1: Comparison against prior work.** To motivate our approach in using language models to scale bibliometric analysis, we compare the scale of our analysis against prior work looking at gender disparities in academic medical research. Our approach enables us to analyze an order of magnitude more authors, manuscripts, and journals to gain insights into research trends in academic medicine compared with existing work. Furthermore, our proposed method achieves an annotation success rate that is at least that of other automated methods.

| Medical Specialty | Broad Subject Terms |
|---|---|
| Medicine | Internal Medicine<br>Medicine – includes General Medicine, Medical Research, Acta Medica and State Medical Journals |
| Allergy | Allergy and Immunology – includes Hypersensitivity, Lymphology, Serology, Serotherapy, and Interferons |
| Cardiology | Cardiology<br>Vascular Diseases – includes Blood Circulation, Hypertension, and Thrombosis |
| Critical Care | Critical Care – includes Intensive Care |
| Endocrinology | Endocrinology – includes Hormones and Diabetes Mellitus<br>Metabolism – includes Obesity |
| Gastroenterology | Gastroenterology – includes Diseases of the Digestive System (Esophagus, Liver, Gallbladder, and Pancreas) |
| Geriatrics | Geriatrics – includes Aging and the Aged |
| Infectious Disease | Acquired Immunodeficiency Syndrome<br>Anti-Infective Agents – includes Antibacterial, Antifungal, Antiviral, and Antiparasitic Agents<br>Bacteriology<br>Parasitology – includes Protozoology<br>Virology |
| Oncology | Antineoplastic Agents<br>Neoplasms – includes Medical and Experimental Oncology |
| Nephrology | Nephrology – includes Dialysis and Hemodialysis |
| Primary Care | Primary Health Care – includes Family Practice |
| Pulmonology | Pulmonary Medicine – includes Respiratory Tract Diseases, Thoracic Diseases, and Pulmonary Tuberculosis |
| Rheumatology | Rheumatology – includes Arthritis |

**Supplementary Table 2: National Library of Medicine (NLM) Broad Subject Terms included in each medical specialty.** In our work, we define a set of 13 medical specialties that include academic journals associated with specific Broad Subject Terms for Indexed Journals made available by the NLM. Broad Subject Terms "are assigned by NLM to MEDLINE journals to describe the journal's overall scope" according to the NLM.[9]

| | Reference Annotation Source | | |
|---|---|---|---|
| **Gender Annotation Method** | **Human Annotator 1** | **Human Annotator 2** | **Human Annotators (Consensus)** |
| SexMachine[10] | 0.337 | 0.438 | 0.339 |
| GenderizeR[11] | 0.473 | 0.499 | 0.516 |
| Global Gender Predictor[12] | 0.873 | 0.913 | 0.930 |
| Database Consensus | 0.485 | 0.548 | 0.526 |
| GenderAPI[13] | 0.870 | 0.923 | 0.945 |
| Meta Llama-3.1 8B[14] (Single) | 0.878 ± 0.007 | 0.906 ± 0.005 | 0.957 ± 0.004 |
| <u>Meta Llama-3.1 8B (Consensus)</u> | <u>0.924</u> | <u>0.935</u> | <u>0.967</u> |
| OpenAI GPT-OSS 20B[15] (Single) | 0.830 ± 0.008 | 0.903 ± 0.003 | 0.924 ± 0.008 |
| OpenAI GPT-OSS 20B (Consensus) | 0.884 | 0.947 | 0.966 |
| OpenAI GPT-OSS 120B[16] (Single) | 0.669 ± 0.011 | 0.718 ± 0.011 | 0.724 ± 0.007 |
| OpenAI GPT-OSS 120B (Consensus) | 0.842 | 0.889 | 0.916 |
| Human Annotator 1 | 1.000 | 0.945 | 1.000 |
| Human Annotator 2 | 0.945 | 1.000 | 1.000 |
| Human Annotators (Consensus) | 1.000 | 1.000 | 1.000 |

**Supplementary Table 3: Evaluation of the agreement between automated gender annotation strategies and reference human annotations.** We show the Cohen's kappa score that quantifies the inter-rater agreement between the reference human annotations and each of the automated gender annotation strategies. "Consensus" refers to the threshold-based algorithm of deriving a consensus gender label from multiple independent annotations, as described in the **Predicting the Gender of Manuscript Authors Using a Large Language Model** subsection in the **Materials and Methods** in the main text; we report results using $\tau$ = 2 (out of 3) for the database methods and $\tau$ = 4 (out of 5) for the LLM-based methods. "Single" refers to using only a single prediction for data labelling (i.e., ablating the consensus algorithm). Error values represent the standard error of the mean (SEM). The method used in the main text and in our subsequent supplementary experiments is <u>underlined</u>.

| Failure Mode | Frequency (%) | Examples | Description |
|---|---|---|---|
| Out-of-Distribution | 74.0 | Foo Chai<br>新利<br>Nkoke | The first name is under-represented in the US, and so the Llama-3.1 8B LLM from Meta AI (a company in the US) is unlikely to correctly predict the gender. |
| Unisex | 13.1 | Jamie<br>Andy<br>Taylor | The first name is gender-neutral and can be traditionally associated with both females and males. |
| Initials | 11.4 | H. A. W.<br>G R<br>M.A.S. | The first name consists of the author's first initial and possible middle initial(s). |
| String Encoding Error | 1.2 | Ren√©e (Renée)<br>Christian¬†Z (Christian Z)<br>Aim√© (Aimé) | The encoding used by the NLM Entrez API results in garbled text and misrepresentations of the first name. |
| Organization | 0.3 | SIMBA and CoMICs<br>START2-POST-VTE<br>STRATOS | The first name is the name or abbreviation of an organization, and not a singular individual. |

**Supplementary Table 4: Failure modes of the LLM-based gender annotation pipeline.** We describe the observed failure modes of our pipeline (i.e., the "Meta Llama-3.1 8B (Consensus)" pipeline in **Supplementary Table 3** and in **Fig. 1b**) when it failed to assign a gender to a first name for 1000 randomly selected examples from our dataset, report their respective frequencies, and list 3 examples of each failure mode. In the "String Encoding Error" row, we also provide the predicted correct representation of each example name in parentheses; the predicted correct representation was generated by encoding the original name in Mac OS Roman and then decoding it back into UTF-8, recovering the original characters when possible.

| | F/F Count (%) | F/M Count (%) | M/F Count (%) | M/M Count (%) | Total (%) |
|---|---|---|---|---|---|
| Medicine | 86,605 (19.1) | 102,078 (22.5) | 53,830 (11.9) | 130,964 (28.9) | 373,477 (82.5) |
| Allergy | 19,168 (19.7) | 23,563 (24.3) | 11,512 (11.9) | 24,506 (25.2) | 78,749 (81.1) |
| Cardiology | 7,873 (12.2) | 13,531 (21.0) | 6,835 (10.6) | 26,194 (40.6) | 54,433 (84.3) |
| Critical Care | 2,300 (12.8) | 3,656 (20.3) | 1,964 (10.9) | 7,680 (42.6) | 15,600 (86.5) |
| Endocrinology | 8,059 (20.1) | 9,533 (23.8) | 4,343 (10.9) | 10,326 (25.8) | 32,261 (80.6) |
| Gastroenterology | 5,992 (14.1) | 9,379 (22.1) | 4,378 (10.3) | 14,980 (35.2) | 34,729 (81.7) |
| Geriatrics | 10,425 (28.6) | 8,528 (23.4) | 4,595 (12.6) | 7,485 (20.5) | 31,033 (85.1) |
| Infectious Disease | 6,416 (17.5) | 9,807 (26.7) | 4,288 (11.7) | 10,013 (27.3) | 30,524 (83.2) |
| Nephrology | 3,833 (16.7) | 5,467 (23.8) | 2,417 (10.5) | 7,879 (34.3) | 19,596 (85.4) |
| Oncology | 36,733 (18.4) | 43,893 (21.9) | 22,292 (11.1) | 55,597 (27.8) | 158,515 (79.2) |
| Primary Care | 6,147 (32.2) | 3,721 (19.5) | 2,268 (11.9) | 5,102 (26.7) | 17,238 (90.2) |
| Pulmonology | 6,578 (14.3) | 9,597 (20.8) | 4,911 (10.6) | 16,895 (36.6) | 37,981 (82.3) |
| Rheumatology | 3,818 (22.9) | 3,814 (22.8) | 2,108 (12.6) | 4,276 (25.6) | 14,016 (83.9) |
| **Total** | **203,947 (18.7)** | **246,567 (22.6)** | **125,741 (11.5)** | **321,897 (29.4)** | **898,152 (82.2)** |

**Supplementary Table 5: Distribution of manuscripts stratified by predicted genders of first and last authors.** We stratify the total number of manuscripts in our dataset based on the LLM-predicted genders of the first and last authors. Percentages are with respect to the total number of manuscripts—including those where either the gender of the first and/or last author could not be determined. The column "**[First]/[Last] Count**" refers to the number of manuscripts published in the journal with a predicted first author gender of [First] and last author gender of [Last], where **M** corresponds to male and **F** to female. Note that the percentages do not necessarily sum to 100% in the **Total Count** column, as some manuscripts exist where the gender of the first and/or last author could not be determined.

| | Journal | SJR (2024) | Manuscript Count (%) | Author Count (%) | |
|---|---|---|---|---|---|
| | | | | Female | Male |
| Medicine | PloS One | 0.80 | 90,997 (20.1) | 279,132 (39.2) | 372,084 (52.3) |
| | Medicine | 0.47 | 50,284 (11.1) | 247,037 (40.1) | 303,691 (49.3) |
| | BMJ Open | 1.02 | 32,772 (7.24) | 131,269 (48.6) | 115,914 (42.9) |
| Allergy | Frontiers in Immunology | 1.94 | 39,868 (41.1) | 142,459 (44.5) | 138,474 (43.3) |
| | Human Vaccines & Immunotherapeutics | 0.90 | 4,914 (5.06) | 14,833 (45.3) | 13,821 (42.2) |
| | Infection and Immunity | 1.05 | 3,561 (3.67) | 11,350 (42.7) | 12,845 (48.4) |
| Cardiology | Journal of the American Heart Association | 2.19 | 9,409 (14.6) | 36,154 (32.2) | 63,399 (56.5) |
| | BMC Cardiovascular Disorders | 0.71 | 4,859 (7.53) | 12,797 (35.4) | 17,364 (48.0) |
| | Cardiovascular Diabetology | 3.35 | 2,610 (4.04) | 9,795 (36.9) | 13,270 (50.0) |
| Critical Care | Critical Care | 2.74 | 5,192 (28.8) | 26,725 (30.9) | 43,786 (50.6) |
| | American Journal of Respiratory and Critical Care Medicine | 5.08 | 4,155 (23.0) | 10,965 (35.3) | 18,613 (60.0) |
| | Critical Care Medicine | 2.12 | 1,379 (7.65) | 3,885 (34.8) | 6,747 (60.4) |
| Endocrin-ology | Oxidative Medicine and Cellular Longevity | 1.67 | 5,768 (14.4) | 18,443 (42.8) | 16,996 (39.4) |
| | Redox Biology | 3.37 | 3,158 (7.89) | 12,183 (41.7) | 12,598 (43.2) |
| | Journal of Clinical Endocrinology & Metabolism | 2.18 | 3,010 (7.52) | 14,666 (48.6) | 13,451 (44.6) |
| Gastro-enterology | World Journal of Gastroenterology | 1.42 | 7,553 (17.8) | 18,351 (36.2) | 25,188 (49.7) |
| | BMC Gastroenterology | 0.84 | 2,337 (5.50) | 12,718 (34.3) | 18,480 (49.8) |
| | Hepatology Communications | 2.11 | 1,813 (4.26) | 7,275 (39.3) | 9,752 (52.7) |
| Geriatrics | Geriatrics | 0.67 | 4,594 (12.6) | 28,952 (50.6) | 21,869 (38.3) |
| | Journal of Alzheimer's Disease | 1.17 | 2,563 (7.03) | 9,863 (45.6) | 10,523 (48.6) |
| | Journal of the American Geriatrics Society | 1.89 | 2,515 (6.90) | 9,823 (54.8) | 7,308 (40.7) |
| Infectious Disease | Viruses | 1.15 | 14,659 (40.0) | 52,225 (45.5) | 53,943 (47.0) |
| | Journal of Virology | 1.28 | 7,936 (21.6) | 26,794 (40.2) | 32,450 (48.7) |
| | Virology Journal | 0.99 | 2,412 (6.58) | 8,292 (41.9) | 8,324 (42.1) |
| Nephrology | BMC Nephrology | 0.81 | 4,261 (18.6) | 13,965 (38.8) | 16,975 (47.2) |
| | Clinical Journal of the American Society of Nephrology | 2.38 | 2,709 (11.8) | 6,889 (39.1) | 9,724 (55.2) |
| | Journal of the American Society of Nephrology | 3.75 | 2,682 (11.7) | 9,217 (37.3) | 13,980 (56.6) |
| Oncology | Neuro-oncology | 6.97 | 17,943 (8.97) | 59,423 (35.1) | 83,887 (49.6) |
| | Oncotarget | 0.79 | 16,427 (8.21) | 56,322 (39.0) | 66,949 (46.4) |
| | Cancer | 2.95 | 7,830 (3.91) | 51,528 (39.1) | 60,319 (45.8) |
| Primary Care | British Journal of General Practice | 1.37 | 4,171 (21.8) | 6,371 (47.0) | 6,509 (48.0) |
| | Annals of Family Medicine | 1.11 | 3,218 (16.8) | 9,448 (59.2) | 5,827 (36.5) |
| | Atencion Primaria | 0.43 | 1,440 (7.54) | 3,220 (53.6) | 2,591 (43.1) |
| Pulmon-ology | BMC Pulmonary Medicine | 0.89 | 3,682 (7.98) | 11,289 (37.8) | 14,143 (47.3) |
| | Journal of Cardiothoracic Surgery | 0.49 | 3,540 (7.67) | 5,625 (26.1) | 12,626 (58.5) |
| | Thoracic Cancer | 0.80 | 3,059 (6.63) | 8,057 (31.2) | 13,253 (51.3) |
| Rheumat-ology | Rheumatology | 1.72 | 5,332 (31.9) | 39,733 (37.8) | 52,482 (49.9) |
| | Arthritis Research & Therapy | 1.59 | 2,769 (16.6) | 11,368 (40.9) | 13,571 (48.9) |
| | Pediatric Rheumatology Online Journal | 0.91 | 1,328 (7.95) | 9,530 (41.8) | 6,858 (30.1) |

**Supplementary Table 6: High-volume journals by medical subspecialty.** We report the 3 journals with the greatest number of unique manuscripts published between January 2015 and September 2025 within each specialty. The "Manuscript Count" percentage is the percentage of manuscripts published by the journal within the specialty. The "Author Count" percentages are the percentages of female and male authors publishing within each journal; note that they do not necessarily sum to 100% in each row, as the gender of some manuscript authors could not be determined.

| | Journal | SJR (2024) | Manuscript Count (%) | Author Count (%) | |
|---|---|---|---|---|---|
| | | | | Female | Male |
| Medicine | New England Journal of Medicine | 19.1 | 1,618 (0.36) | 9,809 (35.2) | 12,362 (44.3) |
| | Nature Medicine | 18.3 | 1,709 (0.38) | 20,296 (40.5) | 26,500 (52.9) |
| | Nature Reviews Disease Primers | 18.0 | 97 (0.02) | 285 (38.1) | 440 (58.7) |
| Allergy | Nature Reviews Immunology | 17.5 | 463 (0.48) | 495 (37.2) | 771 (57.9) |
| | Annual Review of Immunology | 16.4 | 75 (0.08) | 99 (41.3) | 137 (57.1) |
| | Immunity | 12.2 | 1,161 (1.20) | 6,261 (38.5) | 8,715 (53.6) |
| Cardiology | Circulation | 8.67 | 2,290 (3.55) | 9,527 (32.4) | 18,156 (61.7) |
| | JACC: Cardiovascular Imaging | 5.43 | 393 (0.61) | 1,642 (28.9) | 3,772 (66.5) |
| | Circulation Research | 4.90 | 1,838 (2.85) | 5,517 (36.4) | 8,467 (55.9) |
| Critical Care | American Journal of Respiratory and Critical Care Medicine | 5.08 | 4,155 (23.0) | 10,965 (35.3) | 18,613 (60.0) |
| | Intensive Care Medicine | 5.00 | 1,179 (6.54) | 12,770 (38.0) | 17,554 (52.2) |
| | Critical Care | 2.74 | 5,192 (28.8) | 26,725 (30.9) | 43,786 (50.6) |
| Endocrin-ology | Cell Metabolism | 11.9 | 994 (2.48) | 4,857 (38.0) | 6,864 (53.8) |
| | Nature Metabolism | 7.53 | 514 (1.28) | 3,389 (39.7) | 4,506 (52.8) |
| | Bone Research | 4.05 | 452 (1.13) | 1,790 (38.4) | 2,003 (42.9) |
| Gastro-enterology | The Lancet Gastroenterology & Hepatology | 11.4 | 237 (0.56) | 2,359 (38.3) | 3,410 (55.4) |
| | Journal of Hepatology | 10.6 | 687 (1.62) | 3,349 (36.7) | 5,135 (56.3) |
| | Nature Reviews Gastroenterology and Hepatology | 9.91 | 214 (0.50) | 321 (33.1) | 594 (61.2) |
| Geriatrics | Nature Aging | 7.08 | 331 (0.91) | 2,149 (39.0) | 2,578 (46.7) |
| | The Lancet Healthy Longevity | 5.17 | 173 (0.47) | 1,010 (42.2) | 1,137 (47.5) |
| | Ageing Research Reviews | 4.22 | 212 (0.58) | 440 (37.4) | 622 (52.9) |
| Infectious Disease | Nature Microbiology | 6.89 | 903 (2.46) | 5,696 (39.9) | 7,600 (53.3) |
| | The Lancet Microbe | 5.26 | 430 (1.17) | 2,968 (43.5) | 3,282 (48.1) |
| | Annual Review of Virology | 4.03 | 74 (0.20) | 97 (39.9) | 119 (49.0) |
| Nephrology | Nature Reviews Nephrology | 7.31 | 251 (1.09) | 446 (36.1) | 745 (60.2) |
| | Kidney International | 4.13 | 832 (3.63) | 3,865 (38.0) | 5,511 (54.2) |
| | Journal of the American Society of Nephrology | 3.75 | 2,682 (11.7) | 9,217 (37.3) | 13,980 (56.6) |
| Oncology | CA: A Cancer Journal for Clinicians | 145 | 98 (0.05) | 423 (48.8) | 420 (48.5) |
| | Nature Reviews Clinical Oncology | 28.7 | 180 (0.09) | 387 (35.9) | 651 (60.4) |
| | Nature Reviews Cancer | 24.4 | 208 (0.10) | 300 (32.3) | 577 (62.2) |
| Primary Care | British Journal of General Practice | 1.37 | 4,171 (21.8) | 6,371 (47.0) | 6,509 (48.0) |
| | Family Process | 1.33 | 248 (1.30) | 666 (61.2) | 353 (32.4) |
| | Family Medicine and Community Health | 1.15 | 237 (1.24) | 719 (50.8) | 610 (43.1) |
| Pulmon-ology | Journal of Thoracic Oncology | 7.95 | 578 (1.25) | 2,460 (34.5) | 3,477 (48.7) |
| | The Lancet Respiratory Medicine | 6.78 | 744 (1.61) | 5,608 (31.1) | 8,021 (44.6) |
| | The European Respiratory Journal | 4.50 | 1,333 (2.89) | 6,316 (37.1) | 9,608 (56.5) |
| Rheumat-ology | Nature Reviews Rheumatology | 5.51 | 160 (0.96) | 304 (41.2) | 402 (54.5) |
| | The Lancet Rheumatology | 3.84 | 340 (2.03) | 3,572 (45.8) | 3,781 (48.5) |
| | Arthritis and Rheumatology | 3.78 | 1,277 (7.64) | 7,445 (42.4) | 8,235 (46.8) |

**Supplementary Table 7: High-impact journals by medical subspecialty.** We report the 3 journals with the highest 2024 SCImago Journal Rank (SJR, a measure of journal impact) score within each specialty. The “Manuscript Count” percentage is the percentage of manuscripts published by the journal within the specialty. The “Author Count” percentages are the percentages of female and male authors publishing within each journal; note that they do not necessarily sum to 100% in each row, as the gender of some manuscript authors could not be determined.

| Medical Specialty | Single-Author Manuscript Count (%) | Single-Author Manuscript Counts by Gender (%) | |
|---|---|---|---|
| | | Female | Male |
| Medicine | 18,479 (3.80) | 5,667 (30.7) | 10,350 (56.0) |
| Allergy | 2,741 (2.61) | 891 (32.5) | 1,686 (61.5) |
| Cardiology | 2,374 (3.52) | 475 (20.0) | 1,773 (74.7) |
| Critical Care | 1,088 (5.84) | 236 (21.7) | 776 (71.3) |
| Endocrinology | 1,193 (2.74) | 243 (20.4) | 652 (54.7) |
| Gastroenterology | 1,535 (3.43) | 421 (27.4) | 997 (65.0) |
| Geriatrics | 1,020 (2.64) | 376 (36.9) | 553 (54.2) |
| Infectious Disease | 884 (2.25) | 236 (26.7) | 562 (63.6) |
| Nephrology | 1,251 (5.26) | 407 (32.5) | 797 (63.7) |
| Oncology | 3,816 (1.75) | 1,145 (30.0) | 2,231 (58.5) |
| Primary Care | 3,724 (18.64) | 1,382 (37.1) | 2,162 (58.1) |
| Pulmonology | 1,917 (3.99) | 441 (23.0) | 1,361 (71.0) |
| Rheumatology | 409 (2.31) | 153 (37.4) | 202 (49.4) |
| **Total** | 40,431 (3.45) | 12,073 (29.9) | 24,102 (59.6) |

**Supplementary Table 8: Analysis of single-author manuscripts by medical specialty.** We report the total number of manuscripts authored by a single researcher in each medical field, and further stratify these single-author manuscripts in each row by the predicted gender of the author. Note that the sum of the female- and male- authored manuscripts in each row do not necessarily add up to the total number of single-author manuscripts for that specialty, as the gender of some manuscript authors could not be determined using our LLM-based gender annotation pipeline. The percentage values in the second column correspond to the proportion of total manuscripts in the medical specialty that have a single author. The percentage values in the third and fourth columns correspond to the proportion of single-author manuscripts in the medical specialty authored by female and male authors, respectively.

### Supplementary Methods

#### Large Language Model Prompt

Our large language model (LLM) prompt used to predict the gender of an input name <|NAME|> is shown below. Newline characters are represented by the string \n. For phrase sets in blue text, exactly one phrase is sampled independently and randomly from the set with equal probability for each LLM query. In this way, there are 4 distinct LLM prompts that are used for the gender prediction task; however, note that these prompts are semantically invariant and only involved changing the order of the output options and the in-context examples.

```
Given a name, respond with whether the person is {male or female, female or
male}.\n\n{Alice: female\n\nBob: male, Adam: male\n\nBella: female}\n\n<|NAME|>:
```

#### Supplementary Results

##### Evaluating Alternative Gender Annotation Strategies

In our **Materials and Methods**, we propose a method to use large language model (LLM) to annotate the genders of manuscript authors. However, other gender annotation strategies are also possible and have been used in prior work. To evaluate the efficacy of our LLM-based approach, we compare against database-based approaches that retrieve gender labels from an exhaustive database, and also against GenderAPI, which is a commercial solution that uses proprietary methods to assign predicted gender labels given an input first name. Separately, we also evaluate GPT-OSS 20B (`openai/gpt-oss-20b`) and 120B (`openai/gpt-oss-120b`) models using our LLM-based approach, which are more recently released, state-of-the-art open-source language models compared to Meta Llama-3.1 8B.

Our experimental results comparing each of these methods to the small dataset of reference human annotations described in the main text is shown in **Supplementary Table 3**. We find that our proposed LLM-based approach agrees better with reference human annotations when compared to alternative methods. Interestingly, we found that Meta Llama-3.1 8B subjectively exceeded the performance of the GPT-OSS models; this is likely because our input prompt described in the **Supplementary Methods** was optimized using the Meta Llama-3.1 8B model. Given that Meta Llama-3.1 8B already demonstrated impressive performance and can uniquely be run on existing consumer-grade hardware, we opted to use this model to power our experiments reported in the main text. Common failure modes of our final approach in failing to assign a gender to an author are reported in **Supplementary Table 4**. Future work might explore how to better adapt more powerful models and/or methods for bibliometric analyses.

##### Distribution of Research Journals in Bibliometric Analysis

Traditional bibliometric analysis studies have limited the research scope to manuscripts published in a limited number of journals (**Supplementary Table 1**). In contrast, our work analyzes manuscripts from hundreds of research journals. To better assess the relative contributions of each journal to our overall analysis, we plot the number of manuscripts published per journal in **Supplementary Figure 1**; additional details are included in **Supplementary Data 2**. Within each medical subspecialty, we also list the 3 journals that published the greatest number of manuscripts between January 2015 and September 2025 in **Supplementary Table 6**, and the 3 journals with the highest impact (as estimated by the 2024 SCImago Journal Rank score) in **Supplementary Table 7**.

**Additional Analysis of Gender Impact on Author Lists**

**Supplementary Table 5** stratifies the distribution of manuscripts based on the predicted genders of first and last authors. Cumulatively across all medical subspecialties, we found that the distribution of manuscripts was statistically unlikely to occur by chance, suggesting that first and last authors may preferentially choose to work with other authors based on gender (Pearson's $\chi^2$ test: $\chi^2$ = 28524.851, df = 1, $p$ = 0.0). However, Cramér's V effect size is $\phi$ = 0.178 suggesting that the association is relatively small in magnitude. The observed odds ratio (OR) is 2.117 ± 0.031 (mean ± 99.9% CI), suggesting that last authors have about 2.117 times the odds of being paired with a first author of the same gender compared to with a first author of the opposite gender. Finally, we also computed the conditional probabilities P(First Author = male | Last Author = female) = 0.381, while P(First Author = female | Last Author = male) = 0.434, which suggests that both male and female last authors are more likely to have a first author of the same gender, with this tendency being more pronounced in male first- and last- author collaborations. We further stratify these results by medical specialty in the table below:

| **Specialty** | $\chi^2$ | ***p*-value** | $\phi$ | **OR** | **P(M First \| F Last)** | **P(F First \| M Last)** |
|---|---|---|---|---|---|---|
| Medicine | 11190.287 | 0.0 | 0.173 | 2.064 ± 0.047 | 0.383 | 0.438 |
| Allergy | 1366.684 | $3.65 \times 10^{-299}$ | 0.132 | 1.732 ± 0.087 | 0.375 | 0.490 |
| Cardiology | 1704.858 | 0.0 | 0.177 | 2.230 ± 0.149 | 0.465 | 0.341 |
| Critical Care | 617.530 | $2.58 \times 10^{-136}$ | 0.199 | 2.460 ± 0.316 | 0.461 | 0.323 |
| Endocrinology | 887.536 | $5.03 \times 10^{-195}$ | 0.166 | 2.010 ± 0.162 | 0.350 | 0.480 |
| Gastroenterology | 1095.796 | $2.71 \times 10^{-240}$ | 0.178 | 2.186 ± 0.178 | 0.422 | 0.385 |
| Geriatrics | 850.366 | $6.05 \times 10^{-187}$ | 0.166 | 1.991 ± 0.162 | 0.306 | 0.533 |
| Infectious Disease | 305.385 | $2.21 \times 10^{-68}$ | 0.100 | 1.528 ± 0.127 | 0.401 | 0.495 |
| Nephrology | 707.931 | $5.64 \times 10^{-156}$ | 0.190 | 2.286 ± 0.249 | 0.387 | 0.410 |
| Oncology | 4864.171 | 0.0 | 0.175 | 2.087 ± 0.074 | 0.378 | 0.441 |
| Primary Care | 1677.467 | 0.0 | 0.312 | 3.716 ± 0.422 | 0.270 | 0.422 |
| Pulmonology | 1449.337 | 0.0 | 0.195 | 2.358 ± 0.184 | 0.427 | 0.362 |
| Rheumatology | 411.962 | $1.37 \times 10^{-91}$ | 0.171 | 2.031 ± 0.249 | 0.356 | 0.471 |

**Analysis of the Gender Distribution of Single-Author Manuscripts**

Here, we analyze the gender-based distribution of manuscripts in each medical specialty that were authored by only a single individual. In **Supplementary Table 8**, we found that a total of approximately 3.45% of all manuscripts had a single author. The specialties with the greatest proportions of single-author manuscripts were Primary Care (18.64%), Critical Care (5.84%), and Nephrology (5.26%). In contrast, the fields of Oncology (1.75%), Infectious Disease (2.25%), and Rheumatology (2.31%) had lower proportions of single-author manuscripts.

There was also a statistically significant difference between the joint specialty-gender distribution of single-author manuscripts with that of all manuscript authors (Pearson's $\chi^2$ test: $\chi^2 = 36770.409$, df = 25, $p = 0.0$). While the global Cramér's V effect size was only $\phi = 0.062$, we found there was a profound bias toward single authorship shared by both genders in Primary Care (Adjusted Standardized Residual (ASR) 81.020 for female authors and 156.963 for male authors), and there was a gender-independent bias against single authorship in Oncology (ASR -40.107 for female authors and 30.541 for male authors). We report all ASR values below:

| | **Adjusted Standardized Residuals (ASR) for Single Author Manuscripts** | |
|---|---|---|
| **Specialty** | **Female** | **Male** |
| Medicine | -15.255 | 30.114 |
| Allergy | -15.445 | 3.233 |
| Cardiology | -10.552 | 12.525 |
| Critical Care | -2.981 | 16.587 |
| Endocrinology | -13.779 | 1.300 |
| Gastroenterology | -5.838 | 5.136 |
| Geriatrics | -12.156 | -3.519 |
| Infectious Disease | -12.746 | -0.666 |
| Nephrology | 7.794 | 21.991 |
| Oncology | -40.107 | -30.541 |
| Primary Care | 81.020 | 156.963 |
| Pulmonology | -3.756 | 21.255 |
| Rheumatology | -10.957 | -10.308 |

Of note, we observed that female authors were generally less likely to publish single-author manuscripts: 11 out of the 13 medical specialties were associated with a female ASR of less than -3.75. These results are consistent with prior work reporting under-representation of female researchers in single-author manuscripts across a variety of scientific disciplines.[16]
