## Supplementary Data Descriptions for "Large language model-based evaluation of the impact of gender in medical research"

### Description of Additional Supplementary Files

**File Name**: Supplementary Data 1

**File Description**: Source data for all figures.

**File Name**: Supplementary Data 2

**File Description**: For each medical specialty, we report the journals categorized under the corresponding Broad Subject Term from the National Library of Medicine (NLM), along with the total number of manuscripts published by each journal between January 2015 and September 2025, inclusive. We then further stratify the total manuscript count by the LLM-predicted genders of the first and last authors: the column "[First]/[Last] Count" refers to the number of manuscripts published in the journal with a predicted first-author gender of [First] and last-author gender of [Last], where M corresponds to male and F to female. The column "Other Count" refers to the number of manuscripts published in the journal where the gender of either the first and/or last author could not be determined. We focus on the 3 journals with the greatest number of published manuscripts in our study period for each medical specialty in **Supplementary Table 6**, and on the 3 journals with the highest SCImago Journal Rank (SJR, a measure of journal impact) in **Supplementary Table 7**.
